# Patterns of orthopedic and trauma admissions to a tertiary teaching and referral health facility in Kenya: Chart review

**DOI:** 10.1101/2022.12.05.22283136

**Authors:** Maxwell Philip Omondi, Joseph Cege Mwangi, Fred Chuma Sitati, Herbert Onga’ngo’

## Abstract

Tertiary hospitals in resource-limited countries treat patients referred but in most cases are the first level of care for the vast majority of patients. As a result, the tertiary facility effectively functions as a primary health care facility. The urban phenomenon of widespread self-referral is associated with low rates of formal referral from peripheral health facilities.

**Study Objective:** To determine the patterns of orthopaedic and trauma admissions to Kenyatta National Hospital.

**Methodology:** This was descriptive study design. Sample size was 905 patient charts for 2021

**Findings:** The mean age was 33.8 years (SD 16.5) with range of 1 - 93 years. Majority 66.3% were between 25 – 64 years with those above 65 years being 40 (4.4%). Children 0-14 years comprised 10.9% of the admissions. Of the 905 admissions, 80.7% were accident and trauma-related admissions while 17.1% were non-trauma related admissions. About 50.1% were facility referrals while 49.9% were walk-ins. Majority of admissions were through Accident and Emergency Department 78.1%, Corporate Outpatient Care 14.9% and Clinic 7.0%. About 78.7% were emergency admissions while 20.8% were elective admissions. Approximately admissions 48.5% were due to Road Traffic Accidents, 20.9% due to falls and non-trauma related conditions represented 17.1%. Close to 44.8% were casual workers and 20.2% unemployed. Education level was also reviewed with 34.0% having primary education and 35.0% having secondary education. A significant proportion of female admissions (33.2%) were due to non-trauma conditions as compared to male admissions (12.8%) (p<0.001). Aadmissions for those aged 25 – 64 years were 3.5 more likely to have emergency admission as compared to those aged 0 – 14 years. Male were 65.1% less likely to have elective admissions compared to female (p<0.001). Those unemployed were 3.9 more likely to have emergency admission compared to businessmen/women. Vast majority (89.2%) of admissions were within Nairobi Metropolitan region

## Introduction

Approximately 90% of the estimated traumatic injuries occur in low and middle-income countries according to World Health Organization (WHO) (1) and this represents an important global public health problem now and in the coming years (2, 3). Admission patterns for trauma cases have shown gender bias with men affected more than females. The young population age-group less than 40 years are affected as compared to the older population with the most common injury mechanism being road traffic accidents, falls, assaults.

Tertiary hospitals in resource-limited countries treat patients referred but in most cases are the first level of care for the vast majority of patients (4). One of the challenges in health care delivery in resource-limited settings is inappropriate utilization of tertiary health facilities that results in patients’ congestion in referral hospitals with simple conditions that can be effectively managed at the lower peripheral health facilities. The majority of these patients are self-referred, bypassing lower-level health facilities in the process (5-8). A study done in Lusaka demonstrated how University Teaching Hospital is bypassed and as a result the tertiary facility effectively functions as a primary health care facility. The urban phenomenon of widespread self-referral is associated with low rates of formal referral from peripheral health facilities (7, 9).

However, the paucity of data regarding the patterns of orthopaedic and trauma admissions in developing countries and particularly sub-Saharan Africa. The purpose of this study is to determine patterns of orthopaedic and trauma admissions to Kenyatta National Hospital.

## Materials and Methods

### Study design

This was a descriptive study design. **Study area:** Kenyatta National Hospital (KNH) Orthopaedic Wards. KNH is the national teaching and referral hospital based in Upperhill, Nairobi, the Capital city of Kenya. It is located along Hospital Road, about 5km from the city center. It has a bed capacity of approximately 2000 beds. **Study period:** 1^st^ February to 31^st^ December 2021 **Study population:** Orthopaedic and trauma inpatient caseload.

### Inclusion criteria

All orthopaedic admissions to KNH during the study period

### Sample size

A total 0f 905 charts were reviewed.

### Sampling procedure

Stratified sampling technique was used. Strata was based on the point of admission: Accident and Emergency (A&E), Orthopaedic Clinic and Corporate Outpatient Centre (COC). The Population Proportion to Size (PPS) was then used to determine the sample for each stratum based on monthly admissions (Table 1). Within each strata systematic sampling was used to sample the charts for review based on the total monthly admissions per point of admission.

**Table 1:** Orthopaedic admissions to KNH stratified by point of admission, 2021

| Month of the year, 2021` | Point of admission |  |  |  |
| --- | --- | --- | --- | --- |
|  | A&E | Clinic | COC | Total |
| February | 94 | 10 | 9 | 113 |
| March | 68 | 4 | 9 | 81 |
| April | 79 | 3 | 10 | 92 |
| May | 67 | 3 | 11 | 81 |
| June | 78 | 5 | 9 | 92 |
| Total | 386 | 25 | 48 | 459 |
| August | 62 | 10 | 15 | 87 |
| September | 66 | 8 | 14 | 88 |
| October | 82 | 8 | 12 | 102 |
| November | 45 | 6 | 27 | 78 |
| December | 66 | 6 | 19 | 91 |
| Total | 321 | 38 | 87 | 446 |

### Recruitment and consenting procedures

The orthopedic and trauma admissions were identified from the a) admission desk of Health Information System at KNH Accident and Emergency Unit b) KNH Orthopedic Outpatient clinic 5 records c) Corporate Outpatient Center.

### Variables

**a)** Socio-demographic characteristics – age, sex, marital status, religion, current residence, Sub-County of residence, County of residence, country of residence, education level, area of accident b) Nature of admission; c) Occupation; d) Type of admission; e) Level of a facility; f) Diagnoses; g) Mode of payment; h) Admission date; i) Mechanisms of injury

### Quality assurance & quality control procedures

A pilot study was conducted during the design of the study protocol to test the data collection tools for relevance, appropriateness to answer the research questions and adjustments of the data collection tools made as necessary. The PI conducted daily review of all the abstracted forms, verified for accuracy, completeness, and compliance to the research protocol.

### Ethical considerations

UoN/KNH Ethics and Research Committee granted ethical approval (ERC No: P852/10/2021). Written informed consent was obtained from parent/guardian of each participant under 18 years of age. Administrative approval was also sought from KNH Medical Research Department and KNH Orthopaedics Department.

### Data management, analysis

The data were entered into a password-protected Redcap database kept by the KNH Medical Research Department. The quantitative data was analyzed using SPSS version 21. Patterns of orthopedic and trauma admissions were determined using descriptive statistics such as frequencies, measures of central tendencies, measures of dispersions while inferential statistics will be calculated using Pearson’s chi-squared tests and Logistic regressions.

### Study limitations

a) Missing and incomplete data – this was be minimized by making phone calls to patients and or relatives/guardians to fill in the missing information. The patients and relatives/guardians contacts were retrieved from patients’ charts b) Effect of COVID 19 pandemic on referrals of cases from peripheral health facilities and walk-in patients – this was addressed by ensuring the data collection period covered the covid period.

## Results

### Basic Profile of the sample population

The overall mean age was 33.8 years (SD 16.5) with range of 1 - 93 years. Majority 600 (66.3%) were between 25 – 64 years with those above 65 years being 40 (4.4%). Children 0-14 years comprised 99 (10.9%) of the orthopaedic and traumatic admissions. Of the 905 orthopaedic admissions, 743 (82.1%) were Accident/trauma-related admissions while 155 (17.1%) were non-trauma related admissions. About 453 (50.1%) were facility referrals while 452 (49.9%) were walk-ins. Majority of orthopaedic and trauma admissions were through Accident and Emergency Department 707 (78.1%), COC 135 (14.9%) and Clinic 63 (7.0%). With regard to the type of admission 712 (78.7%) were emergency admissions while 188 (20.8%) were elective admissions. With respect to the mechanism of injury, 439 (48.5%) were due to Road Traffic Accidents, 189 (20.9%) were due to falls and non-trauma related conditions 155 (17.1%) (Table 2).

**Table 2:**
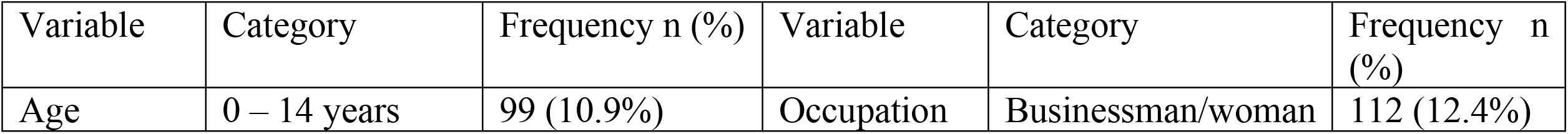
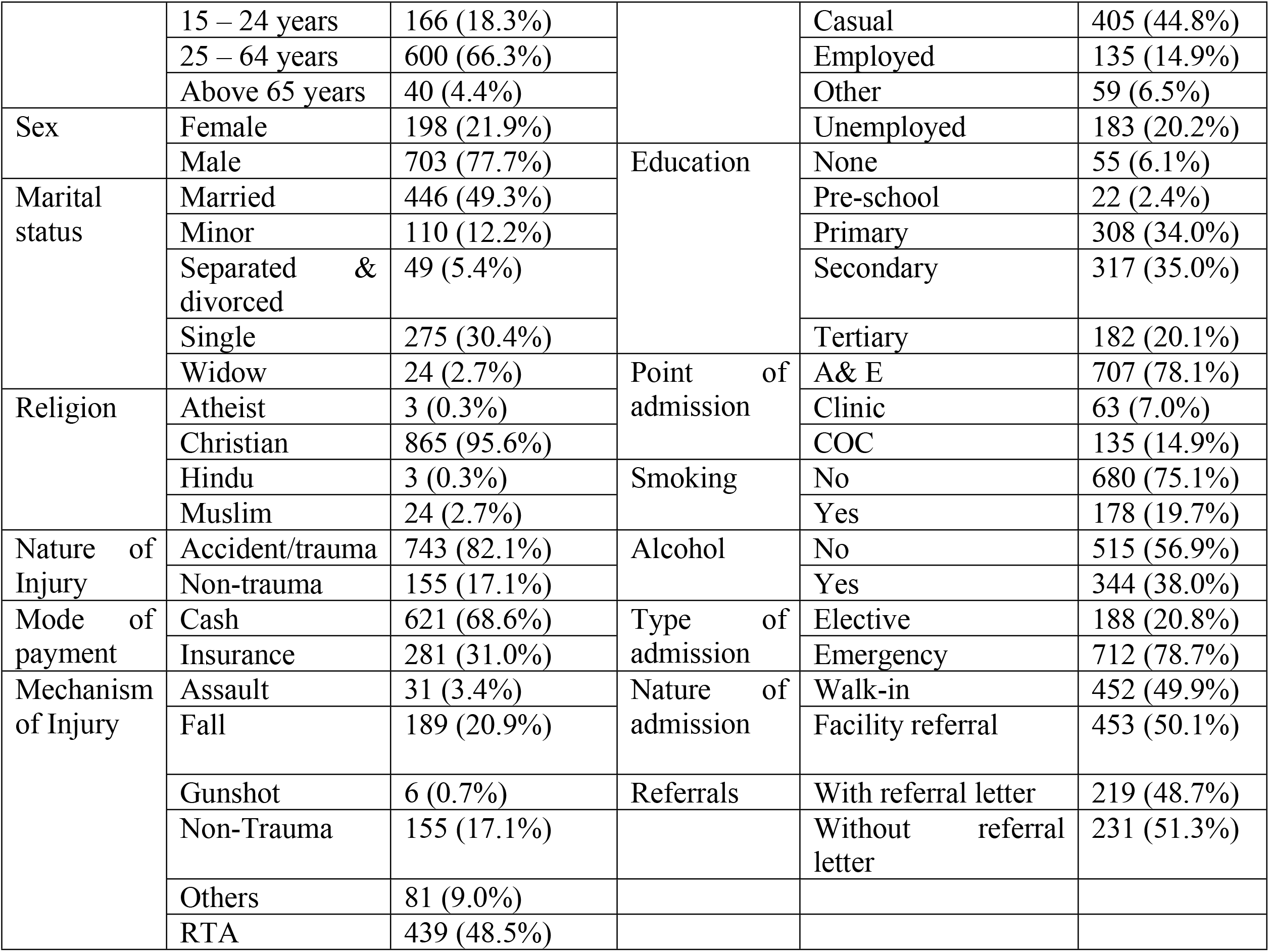
Basic profile of the sample population

About 446 (49.3%) were married, 275 (30.4%) were single and 110 (12.2%) were minor. About 703 (77.7%) were male and 198 (21.9% were female. Christians were the majority at 865 (95.6%) of the orthopaedic and trauma admissions. About 405 (44.8%) were casual workers, 183 (20.2%) unemployed. Education level was also reviewed with 308 (34.0%) having primary education and 317 (35.0%) having secondary education (Table 2).

The study also reviewed the behavioural profile, 178 (19.7%) were smokers and 344 (38.0%) drunk alcohol. With regard to the mode of payment majority 621 (68.6%) were cash payers and 281 (31.0%) had active insurance cover (Table 2).

There were three points for orthopaedic and trauma admissions at KNH namely Accident and Emergency (A&E), Orthopaedic Clinic (Clinic) and Corporate Outpatient Clinic (COC). The study determined the association between the key sociodemographic characteristics (age, sex and education) and the point of admission (A&E, Clinic and COC). Majority of orthopaedic and trauma admissions were through A&E regardless of the age category with significant admissions for those above 65 years of age being admitted through COC (p<0.001) (Table 3). While it was noted that a higher proportion of male 574 (81.9%) and females (127 (64.1%) were admitted through A&E, proportionately more females were admitted through COC (26.3%) (p<0.001) (Table 3). The study also revealed that orthopaedic and trauma admissions for those with secondary level of education and below were majorly admitted through the A&E. For those with tertiary level of education, a significant proportion were admitted through the COC (41.8%) (p<0.0001) (Table 3).

**Table 3:** Association between key socio-demographic characteristics and point of admission

| Variable | Category | Point of admission |  |  | Chi-square; p-value |
| --- | --- | --- | --- | --- | --- |
|  |  | A&E | Clinic | COC |  |
| Age | 0 – 14 years | 72 (72.7%) | 15 (15.2%) | 12 (12.1%) | 27.938; $p < 0.001$ |
|  | 15 – 24 years | 147 (88.6%) | 7 (4.2%) | 12 (7.2%) |  |
|  | 25 – 64 years | 460 (76.9%) | 38 (6.4%) | 100 (16.7%) |  |
|  | Above 65 years | 26 (65.0%) | 3 (7.5%) | 11 (27.5%) |  |
| Sex | Female | 127 (64.1%) | 19 (9.6%) | 52 (26.3%) | 30.045; $p < 0.001$ |
|  | Male | 574 (81.9%) | 44 (6.3%) | 83 (11.8%) |  |
| Education | None | 44 (80.0%) | 5 (9.1%) | 6 (10.9%) | 138.593; $p < 0.001$ |
|  | Pre-school | 21 (95.5%) | 1 (4.5%) | 0 (0.0%) |  |
|  | Primary | 262 (85.3%) | 23 (7.5%) | 22 (7.2%) |  |
|  | Secondary | 264 (83.5%) | 27 (8.5%) | 25 (7.9%) |  |
|  | Tertiary | 100 (54.9%) | 6 (3.3%) | 76 (41.8%) |  |

The study determined the association between the nature of admission and the key socio-demographic variables (age, sex, occupation and education). Nature of admission refers to whether the patient was a walk-in patient or a facility referral. There was no statistically significant association between age, sex, occupation, education and the nature of admission (p>0.05) (Table 4).

**Table 4:** Association between key socio-demographic characteristics and nature of admission

| Variable | Category | Nature of admission |  | Chi-square, p-value |
| --- | --- | --- | --- | --- |
|  |  | Walk-in | Facility Referral |  |
| Age | 0 – 14 years | 38 (38.4%) | 61 (61.6%) | 6.295; $p = 0.098$ |
|  | 15 – 24 years | 87 (52.4%) | 79 (47.6%) |  |
|  | 25 – 64 years | 305 (50.8%) | 295 (49.2%) |  |
|  | Above 65 years | 22 (55.0%) | 18 (45.0%) |  |
| Sex | Female | 111 (56.1%) | 87 (43.9%) | 0.056; $p = 0.056$ |
|  | Male | 340 (48.4%) | 363 (51.6%) |  |
| Occupation | Businessman/woman | 58 (51.8%) | 54 (48.2%) | 1.754; $p = 0.781$ |
|  | Casual | 200 (49.4%) | 205 (50.6%) |  |
|  | Employed | 72 (53.3%) | 63 (46.7%) |  |
|  | Other | 26 (44.1%) | 33 (55.9%) |  |
|  | Unemployed | 89 (48.6%) | 94 (51.4%) |  |
| Education | None | 28 (50.9%) | 27 (49.1%) | 6.886; $p = 0.142$ |
|  | Pre-school | 7 (31.8%) | 15 (68.2%) |  |
|  | Primary | 142 (46.1%) | 166 (53.9%) |  |
|  | Secondary | 159 (50.2%) | 158 (49.8%) |  |
|  | Tertiary | 101 (55.5%) | 81 (44.5%) |  |

The study revealed that a significant proportion of those aged 0 – 14 years (44.9%) and above 65 years (37.5%) were due to falls whereas those between 15 – 24 years (62.7%) and 25 – 64 years (51.4%) were due to RTA (p<0.001) (Table 4). A significant proportion of female admissions (33.2%) were due to non-trauma conditions as compared to male admissions (12.8%) (p<0.001). Road Traffic Accident (RTA) still remains a major mechanism of injury for the KNH admissions (Table 5).

**Table 5:** Association between the socio-demographic characteristics and the mechanism of injury

|  |  | Mechanism of injury |  |  |  |  |  | Chi-square;p-value |
| --- | --- | --- | --- | --- | --- | --- | --- | --- |
| Variable | Category | Assault | fall | gunshot | Non-trauma | Others | RTA |  |
| Age | 0 – 14 years | 1 (1.0%) | 44 (44.9%) | 0 (0.0%) | 18 (18.4%) | 15 (15.3%) | 20 (20.4%) | 91.598; $p<0.001$ |
|  | 15 – 24 years | 6 (3.6%) | 22 (13.3%) | 2 (1.2%) | 16 (9.6%) | 16 (9.6%) | 104 (62.7%) |  |
|  | 25 – 64 years | 24 (4.0%) | 108 (18.1%) | 4 (0.7%) | 108 (18.1%) | 46 (7.7%) | 307 (51.4%) |  |
|  | Above 65 years | 0 (0.0%) | 15 (37.5%) | 0 (0.0%) | 13 (32.5%) | 4 (10.0%) | 8 (20.0%) |  |
| Sex | Female | 1 (0.5%) | 49 (25.0%) | 0 (0.0%) | 65 (33.2%) | 16 (8.2%) | 65 (33.2%) | 59.162; $p<0.001$ |
|  | Male | 30 (4.3%) | 139 (19.8%) | 6 (0.9%) | 90 (12.8%) | 64 (9.1%) | 372 (53.1%) |  |

The study reviewed the type of admission and its association with key socio-demographic characteristics namely age, sex, occupation and education. Orthopaedic and trauma admissions for those aged 25 – 64 years were 70.9% less likely to have elective admissions as compared to those aged 0 – 14 years (Table 6).

**Table 6:** Association between key socio-demographic characteristics and type of admission

| Variable | Category | Emergency | Elective |  | Logistic regression; OR (95% CI) |
| --- | --- | --- | --- | --- | --- |
| Age | 0 – 14 years | 72 (73.5%) | 26 (26.5%) | 13.550;<br>p=0.004 | 1.0 |
|  | 15 – 24 years | 147 (88.6%) | 19 (11.4%) |  | 0.812 (0.360 – 1.835) |
|  | 25 – 64 years | 466 (78.1%) | 131 (21.9%) |  | 0.291 (0.127 – 0.668) |
|  | Above 65 years | 27 (69.2%) | 12 (30.8%) |  | 0.633 (0.312 – 1.283) |
| Sex | Female | 125 (63.8%) | 71 (36.2%) | 35.813;<br>p<0.001 | 1.0 |
|  | Male | 584 (83.4%) | 116 (16.6%) |  | 2.860 (2.009 – 4.070) |
| Occupation | Businessman/woman | 82 (73.2%) | 30 (26.8%) | 66.366;<br>p<0.001 | 1.0 |
|  | Casual | 361 (89.8%) | 41 (10.2%) |  | 0.833 (0.416 – 1.669) |
|  | Employed | 80 (59.3%) | 55 (40.7%) |  | 0.259 (0.136 – 0.491) |
|  | Other | 41 (69.5%) | 18 (30.5%) |  | 0.626 (0.324 – 1.208) |
|  | Unemployed | 142 (78.5%) | 39 (21.5%) |  | 1.566 (0.816 – 3.005) |
| Education | None | 46 (83.6%) | 9 (16.4%) | 55.091;<br>p<0.001 | 1.0 |
|  | Pre-school | 20 (90.9%) | 2 (9.1%) |  | 0.292 (0.135 – 0.633) |
|  | Primary | 260 (85.2%) | 45 (14.8%) |  | 0.149 (0.034 – 0.658) |
|  | Secondary | 264 (83.8%) | 51 (16.2%) |  | 0.258 (0.168 – 0.399) |
|  | Tertiary | 109 (59.9%) | 73 (40.1%) |  | 0.288 (0.189 – 0.440) |

Male were 2.860 (2.009 – 4.070) more likely to have emergency admissions compared to female and this was statistically significant (Table 6). Regarding employment status, those employed were 74.1% less likely to have emergency admissions compared to businessmen/women (Table 6).

Education status was also reviewed and the study showed that those with some level of education were less likely to have emergency admissions compared with those no education and this was statistically significant (Table 6).

The study reviewed the mode of payment and its association with key socio-demographic characteristics namely age, sex, occupation, education, smoking and alcohol intake. The study revealed no statistically significant association between the different age categories (Table 7). Male admissions were 2.003 (1.445 – 2.775) more likely to be cash payers as compared to female (Table 7). His means that female admissions tended to have active insurance cover compared to male admissions.

**Table 7:**
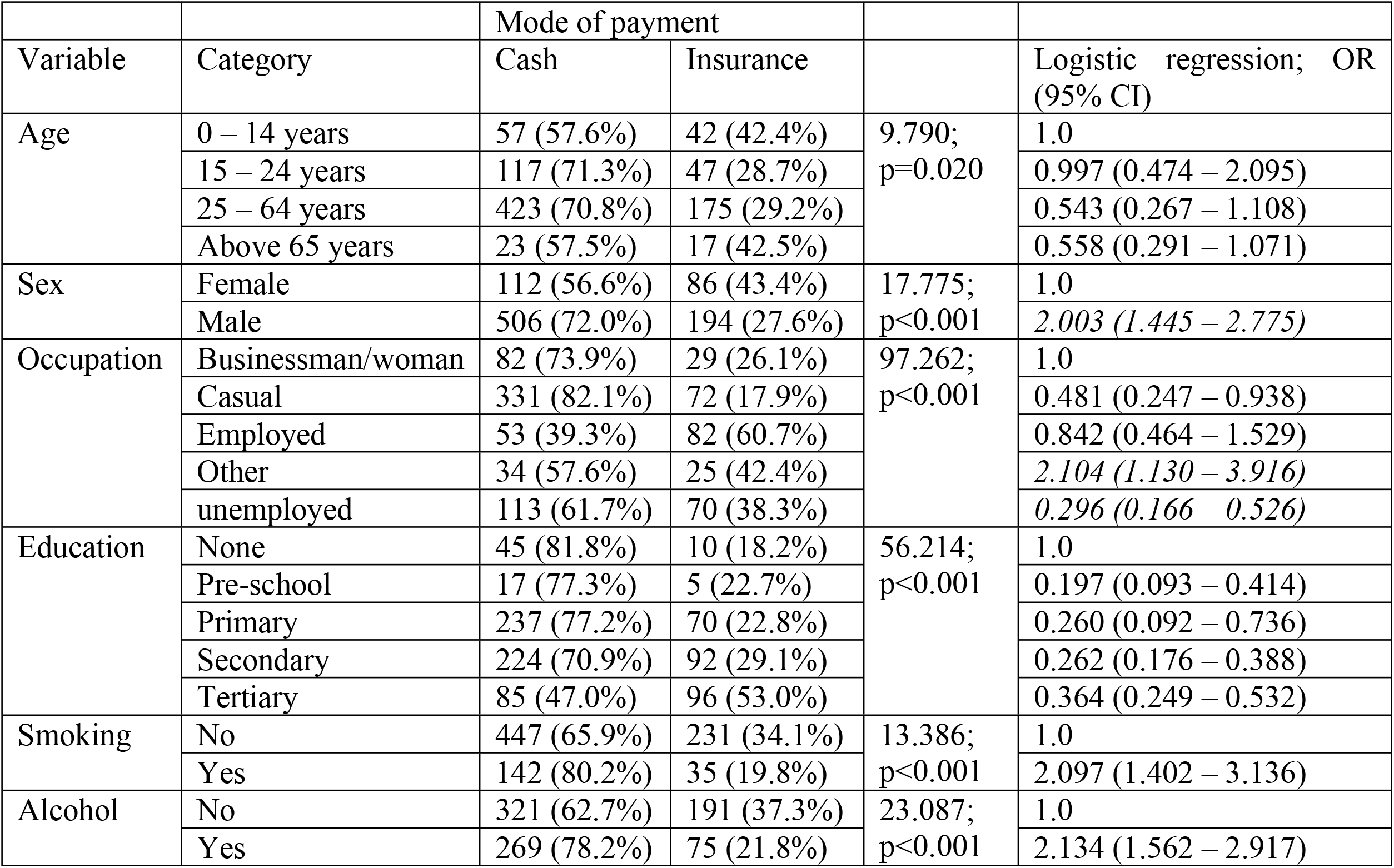
Association between key socio-demographic characteristics and mode of payment

With regard to occupation status, those who are unemployed were 70.4% less likely to be cash payers as compared to businessmen/women (Table 7). Orthopaedic and trauma admissions who were smokers were 2.097 (1.402 – 3.136) more likely to be cash payers as compared to those who did not smoke. In addition, orthopaedic and trauma admissions who took alcohol were 2.134 (1.562 – 2.917) more likely to be cash payers as compared to those who did not take alcohol (Table 7).

The study sought to reveal the patterns of distribution of orthopedic and trauma admissions by sub-county of origin. Most of the admissions were from Nairobi County at 60.3% with 89.2% of patients coming within Nairobi Metropolitan area (a radius of about 40km from Nairobi) Figure 1.

**Figure 1:**
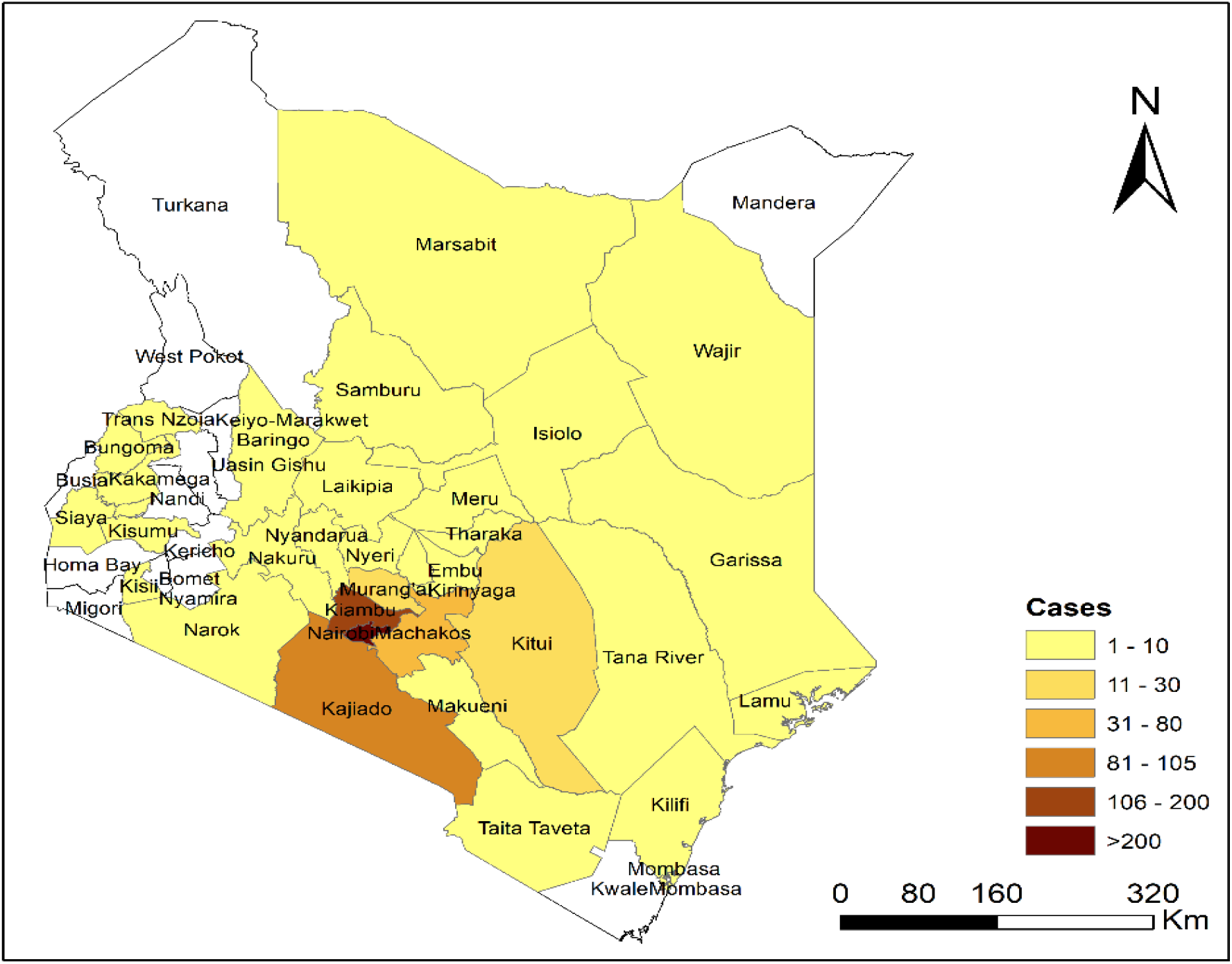
Distribution of orthopedic and trauma admissions to KNH by counties, 2021

## Discussions

It was noted that a higher proportion of orthopaedic and trauma admissions were admitted through A&E and this is in tandem with the finding that about three-fourth off the admissions are emergency. However, a higher proportion of females were admitted through COC which is a private wing of KNH and these tended to be older women above 65 years of age with fragility fractures and musculoskeletal degenerative disorders. These were women with active insurance cover and were of relatively higher economic status.

In addition, the study also revealed that orthopaedic and trauma admissions for those with secondary level of education and below were majorly admitted through the A& E since these were mostly casual workers caught up in road traffic accidents. For those with tertiary level of education, a significant proportion were admitted through the COC and these patients had active insurance cover, better educated and of higher socio-economic status.

The study revealed that about two-thirds of the orthopaedic and trauma admission were either casuals or unemployed. This compares favourably with study done in Taiwan that revealed orthopaedic fractures were associated with patients of low socio-economic status (10). However, it contrasted with studies done on orthopedic admissions in Kilimanjaro Christian Medical Centre (KCMC) in Northern Tanzania showed the three most common occupations were farmers, businessman, professional drivers, students (11, 12). This is could be because KCMC is a private facility as opposed to KNH which is a public health facility. Those who were unemployed were less likely to have insurance cover compared to businessmen and women and this could be attributed to lack of any source of income.

The study revealed the mean age for orthopaedic and trauma admissions were young at 33.816 years with majority between 25 – 64 years. Children and those above 65 years of age were the minority age groups. This compares with studies done in Rwanda, Uganda, Botswana, India, United States of America, Brazil that showed orthopaedic and trauma admissions are of younger age group (13-19).

Majority of orthopaedic and trauma admissions were males at 77.7%. This compares favourably with studies done in Taiwan, India, Botswana, Tanzania, Rwanda, South Africa and Brazil that reveals male predominance (10, 13, 16-18, 20-22). However, this contradicts a retrospective study done in England that revealed approximately equal male to female ratio (20). A significant proportion of female admissions were due to non-trauma conditions as compared to male admissions and this could be due to an increase in female admissions above 65 years of age due to musculoskeletal degenerative disorders. In addition, male orthopaedic admissions predominated and peaked at 25 – 64 years and these admissions declined steadily to 65years of age and orthopaedic admissions were comparable across gender for those above 65 years of age. This compares with a retrospective study done in a tertiary Hospital in Nepal that showed similar admission rates from 60 years of age (23). Male were less likely to have elective admissions compared to female and this is explained by the fact that majority were emergency admissions which males predominated.

About 50.1% were facility referrals while 49.9% were walk-ins. of the facility referrals about 48.7% had written referral letters. This contradicts study done in Meru, Kenya that showed self-referrals accounted for about 80% of referrals to Meru District Hospital (24). This could be due to the fact that the study focussed on non-surgical patients. A similar study done in Kwazulu Natal, South Africa revealed that self-referrals were 36% with 64% being appropriately referred with written referral letter (25). On the other hand a study done in Nigeria showed 92.9% of patients in tertiary hospital directly went to the facility without a referral letters while 7.1% were referred (26). The disparities of these findings could be due to the fact that this study specifically reviewed orthopaedic and trauma admissions and not all medical conditions.

Majority of orthopaedic trauma admissions were due to RTA at 48.5% followed by falls and non-trauma related conditions 17.1%. This compares favourably with studies done in Rift Valley Provincial Hospital in Kenya, Tanzania, Rwanda, India, USA and Taiwan that revealed RTA is the major cause of orthopaedic trauma admissions (10-14, 22, 27, 28). However, this differs with retrospective study done in South Africa that revealed interpersonal violence was the major mechanism of injury of orthopaedic admissions at 60% followed by RTA at 19% (18). Also, a retrospective study done in Botswana and Tanzania showed falls as a leading cause of orthopaedic injuries and RTA second at 36% (21, 28). Falls were the commonest mechanism of injury amongst orthopaedic admissions in children and those above 65 years of age. This compares favourably with similar studies done in Middle East region, Botswana that showed RTA was the major cause of admissions at for younger populations while falls commonest cause of admissions for the extremes of age – children and the elderly (28, 29). This also compares with a multicenter observation study done on distribution of orthopedic fractures in low and middle-income countries revealed about falls (64.5%) was the main mechanism of injury for those 60 years and above (30). Elderly patients who are more prone to fragility fractures and children are more prone to falls.

However, there was no significant difference when it came to facility referrals being accompanied by official written referral letters from the referring facilities to KNH. This is because most of the referrals were verbal over the telephone and once a verbal consensus has been reached the referring health facilities did not see the need of writing an official referral letter.

About a third of the orthopaedic and trauma admissions had active insurance cover. This contrasts with a multicenter observation study done on distribution of orthopedic fractures in low and middle-income countries revealed about 18% of orthopedic admissions in Africa had health insurance cover (30). It also contrasts with a retrospective study done in PCEA Kikuyu Mission Hospital in Kenya showed about 60.82% of orthopedic patients have insurance cover (31). This could be explained by the fact that PCEA Kikuyu Mission Hospital is a private health facility that admits patients with higher socio-economic status compared to KNH which is a public health facility.

Vast majority of patients admitted come from within the Nairobi Metropolitan area consisting of Nairobi, Kajiado, Machakos and Kiambu counties. This compares with a review of orthopedic admissions in KCMC in Northern Tanzania showed 65.7% of the patients came from state of Kilimanjaro where the hospital is located, 12.7% from Arusha,6.4% from Tanga, 5.9% from Manyara and 1.5% from Singida (11).A similar study done in Muhimbili National Hospital in Tanzania showed only 0.8% of admissions were from outside Dar Es Salaam (32). A similar study done in Blantyre in Malawi also revealed majority of referrals come from within the Tertiary facility (33)

## Conclusions

In conclusion, majority of orthopaedic and trauma admissions were younger patients and mostly males. Women tended to have more admissions above 65 years of age mostly through COC. About equal proportions were noted for facility referrals and walk-ins. About half the facility referrals had written referral letters. Majority of orthopaedic trauma admissions were due to RTA followed by falls and non-trauma related conditions. About three-fourth of admissions were emergencies. Those with no education level were mostly associated with emergency admissions compared with those with some level of education. Employed admissions tended to be elective admissions compared to businessmen and women. About a third of the orthopaedic and trauma admissions had active insurance cover. About two-thirds of the admissions were either casuals or unemployed. Vast majority of admissions were from within Nairobi Metropolitan region.

## Data Availability

We have full access of the original raw data and will share the data on request on short notice.

## References

1. Hanche-Olsen TP, Alemu L, Viste A, Wisborg T, Hansen KS. Trauma care in Africa: a status report from Botswana, guided by the World Health Organization’s “Guidelines for Essential Trauma Care”. World J Surg. 2012 Oct;36(10):2371–83.

2. Murray CJL, AD L. The Global Burden of Disease: A Comprehensive Assessment of Mortality and Disability From Diseases Injuries, and Risk Factors in 1990 and Projected to 2020. Journal [serial on the Internet]. 1996 Date.

3. Murray CJL, AD L. Global Health Statistics: A Compendium of Incidence Prevalence and Mortality Estimates for Over 200 Conditions. Journal [serial on the Internet]. 1996 Date.

4. Hensher M, Price M, Adomakoh S. Referral Hospitals. 2006.

5. Holdsworth G, Garner PA, Harphan T. Crowded outpatient departments in city hospitals of developing countries: a case study from Lesotho. Int J Health Plann Manage. 1993 Oct-Dec;8(4):315–24.

6. London L, Bachmann OM. Paediatric utilisation of a teaching hospital and a community health centre. Predictors of level of care used by children from Khayelitsha, Cape Town. S Afr Med J. 1997 Jan;87(1):31–6.

7. Ohara K, Melendez V, Uehara N, Ohi G. Study of a patient referral system in the Republic of Honduras. Health Policy Plan. 1998 Dec;13(4):433–45.

8. Sanders D, Kravitz J, Lewin S, McKee M. Zimbabwe’s hospital referral system: does it work? Health Policy Plan. 1998 Dec;13(4):359–70.

9. Nordberg E, Holmberg S, Kiugu S. Exploring the interface between first and second level of care: referrals in rural Africa. Trop Med Int Health. 1996 Feb;1(1):107–11.

10. Pan RH, Chang NT, Chu D, Hsu KF, Hsu YN, Hsu JC, et al. Epidemiology of orthopedic fractures and other injuries among inpatients admitted due to traffic accidents: a 10-year nationwide survey in Taiwan. ScientificWorldJournal. 2014;2014:637872.

11. Premkumar A, Massawe HH, Mshabaha DJ, R. Foran J, XiaohanYing, Sheth NP. The burden of orthopaedic disease presenting to a referral hospital in northern Tanzania. Global Surgery. 2015;2(1):70–5.

12. William Mack Hardaker, Mubashir Jusabani, Honest Massawe, Anthony Pallangyo, Rogers Temu, Gileard Masenga, et al. The burden of orthopaedic disease presenting to a tertiary referral center in Moshi, Tanzania: a cross-sectional study. Pan African Medical Journal 2022;42:96. 2022;42(96).

13. Jain A, Goyal V, Varma C. Reflection of Epidemiological Impact on Burden of Injury in Tertiary Care Centre, Pre-COVID and COVID Era: “Lockdown, a Good Fortune for Saving Life and Limb”. Indian J Surg. 2020 Oct 24:1–5.

14. Ovadia P, Szewczyk D, Walker K, Abdullah F, Schmidt-Gillespie S, Rabinovici R. Admission patterns of an urban level I trauma center. Am J Med Qual. 2000 Jan-Feb;15(1):9–15.

15. Stonko DP, Dennis BM, Callcut RA, Betzold RD, Smith MC, Medvecz AJ, et al. Identifying temporal patterns in trauma admissions: Informing resource allocation. PLoS One. 2018;13(12):e0207766.

16. Saikiran Velpula, Laxmi Prasanna Gummadi, Nagaraju Vallepu, Bharath Kumar Dasari, Anchuri. SS. Epidemiology of orthopedic trauma admissions in a multispecialty hospital in Warangal-A retrospective study. Clinical Practice. 2019;16(6).

17. Vikas Verma, Sheela Singh, Girish Kumar Singh, Santosh Kumar, Ajay Singh, Kanika Gupta. DISTRIBUTION OF INJURY AND INJURY PATTERNS IN TRAUMA VICTIMS ADMITTED TO THE TRAUMA CENTRE OF CSMMU, LUCKNOW. Indian Journal Of Community Health. 2013;25(1):52–60.

18. Dhaffala A, Longo-Mbenza B, Kingu JH, Peden M, Kafuko-Bwoye A, Clarke M, et al. Demographic profile and epidemiology of injury in Mthatha, South Africa. Afr Health Sci. 2013 Dec;13(4):1144–8.

19. Nathan N. O’Hara, Rodney Mugarura, Gerard P. Slobogean, Bouchard. M. The Orthopaedic Trauma Patient Experience: A Qualitative Case Study of Orthopaedic Trauma Patients in Uganda. PLOS ONE. October 31 2014;9(10).

20. Taylor A, Young. A. Epidemiology of Orthopaedic Trauma Admissions Over One Year in a District General Hospital in England. The open orthopaedics journal. [Journal]. 2015 29 May 9:191–3.

21. Rutta E, Mutasingwa D, Ngallaba SE, Berege ZA. Epidemiology of injury patients at Bugando Medical Centre, Tanzania. East Afr Med J. 2001 Mar;78(3):161–4.

22. Theoneste Nkurunziza, Gabriel Toma, Jackline Odhiambo, Rebecca Maine, Robert Riviello, Neil Gupta, et al. Referral patterns and predictors of referral delays for patients with traumatic injuries in rural Rwanda. Global Surgery. 2016;160(6):1636–44.

23. Mishra BN, Jha A Gupta. Epidemiology of Orthopaedic Admissions at A Teaching Hospital of Eastern Nepal.. Journal of Nobel Medical College. 2017;6(1):56–62.

24. Mwabu GM. Referral systems and health care seeking behavior of patients: An economic analysis. World Development, Elsevier. 1989;17(1):85–91.

25. Pillay I, Mahomed OH. Prevalence and determinants of self referrals to a District-Regional Hospital in KwaZulu Natal, South Africa: a cross sectional study. Pan Afr Med J. 2019;33:4.

26. Akande T. Referral system in Nigeria: Study of a tertiary health facility. Annals of African Medicine. 2004;3(3):130–3.

27. N Masiira-Mukasa, Ombito. BR. Surgical admissions to the Rift Valley Provincial General Hospital, Kenya East Afr Med J 2002;79(7):373–8.

28. Manwana ME, Mokone GG, Kebaetse M T Y. Epidemiology of traumatic orthopaedic injuries at Princess Marina Hospital, Botswana. South African Orthopaedic Journal. 2018 March 2018;17(1):41–6.

29. Chandrashekara CM, George MA, Al-Marboi BSK. Demography of orthopaedic admissions in a secondary care hospital in oman. Brunei International Medical Journal 2013;9(4):236–42.

30. Pouramin P, Li CS, Sprague S, Busse JW, Bhandari M. A multicenter observational study on the distribution of orthopaedic fracture types across 17 low-and middle-income countries. OTA Int. 2019 Sep;2(3):e026.

31. Kihuba E. Epidemiology and medical costs of orthopedic conditions in a tertiary hospital in Kenya; A five-year analysis of admission data. BMJ Yale. 2022.

32. Simba DO, Mbembati NAA, Museru LML, E.K. L. Referral Pattern of Patients Received at the National Referral Hospital: Challenges in Low Income Countries. East African Journal of Public Health. 2008;5(1):6–9.

33. Pittalis C, Brugha R, Bijlmakers L, Mwapasa G, Borgstein E, Gajewski J. Patterns, quality and appropriateness of surgical referrals in Malawi. Trop Med Int Health. 2020 Jul;25(7):824–33.

